# Genome-wide analysis of genetic pleiotropy and causal genes across three age-related ocular disorders

**DOI:** 10.1101/2022.07.15.22277659

**Authors:** Xueming Yao, Hongxi Yang, Han Han, Xuejing Kou, Yuhan Jiang, Menghan Luo, Yao Zhou, Jianhua Wang, Xutong Fan, Xiaohong Wang, Mulin Jun Li, Hua Yan

## Abstract

**Purpose:** Age-related macular degeneration (AMD), cataract, and glaucoma are leading causes of blindness worldwide. Previous genome-wide association studies (GWASs) have revealed a variety of susceptible loci associated with age-related ocular disorders, yet the genetic pleiotropy and causal genes across these diseases remain poorly understood. This study aims to identify genetic pleiotropic genes among AMD, cataract, and glaucoma.

**Methods:** We leveraged large-scale genetic and observational data from ocular disease GWASs and UK Biobank (UKBB) to investigate correlations among these ocular disorders. We undertook meta-analyses with the largest GWAS summary statistics of these ocular disorders to identify pleiotropic loci. We then comprehensively integrated eye-specific gene expression quantitative loci (eQTLs), epigenomic profiling, and 3D genome data to prioritize causal pleiotropic genes. Pathway enrichment analysis and drug repurposing analysis were also conducted.

**Results:** We found significant pairwise genetic correlations and consistent epidemiological associations among AMD, cataract, and glaucoma. Cross-disease meta-analysis uncovered seven pleiotropic loci, three of which were replicated in an additional cohort. Integration of variants in pleiotropic loci and multiple single-cell omics data identified that Müller cells and astrocytes were likely causal cell types underlying ocular comorbidity. After the integration with multi-omics data, 15 causal genes were identified. We found that pleiotropic genes were essential in nerve development and eye pigmentation, and targetable by existing drugs for the treatment of single ocular disorder.

**Conclusions:** These findings will not only facilitate the mechanistic research of ocular comorbidities but also benefit the therapeutic optimization of age-related ocular diseases.

## Introduction

Age-related ocular diseases, such as age-related macular degeneration (AMD), cataract, and glaucoma, are among the leading causes of irreversible blindness and vision impairment in adults aged ≥ 50 years worldwide ^1^. Previous cross-sectional and case-control studies have reported that cataract and glaucoma commonly co-exist and prevalent glaucoma is associated with neovascular AMD ^2, 3^. A clinical study also showed that over 20% of eye samples from the patients listed for cataract surgery have evidence of AMD according to optical coherence tomography imaging ^4^. The findings motivate the exploration of the extent and causal factors for potential comorbidity among these age-related ocular diseases.

Recent genome-wide association studies (GWASs) have identified hundreds of genetic signals that contribute to the complex pathogenesis of AMD ^5, 6^, cataract ^7^, and glaucoma ^8-10^. These and other genetic studies also suggest a high heritability in susceptibility of AMD (∼50%) ^11^, cataract (35-58%) ^12, 13^, and glaucoma (35-65%) ^14, 15^. Nevertheless, the shared genetic architecture among these age-related ocular diseases is largely unknown. A study investigated the genetic pleiotropy among AMD and several eye-related diseases/traits, and found that patients with AMD demonstrate a genetically reduced risk to develop open-angle glaucoma ^16^, which is inconsistent with other findings ^17, 18^. Moreover, no genetic pleiotropy research has been conducted between cataract and other ocular diseases so far. Given the conflicting results and scant knowledge regarding the shared genetic architecture of these age-related ocular disorders, more systematic investigation based on larger and complementary cohorts is needed.

While different levels of genetic correlations or pleiotropic loci can be faithfully estimated among ocular diseases/traits, the pleiotropic genes and causal variants have not been rigorously examined. This is an important issue to clarify, as the shared biological basis of ocular disorders could represent potential efficient intervention targets for the prevention and treatment of ocular comorbidities. Emerging tissue/cell type-specific epigenomic profiles, single-cell omics atlas as well as molecular phenotype quantitative trait loci (QTLs) mapping on certain ocular diseases revealed many causal variants and genes underlying cell type specificity of disease etiology ^19-23^. Thus, integrating large-scale multi-omics and QTL data from eyes with disease pleiotropy information would provide a promising strategy to decipher the genetic causality of ocular comorbidities, especially for the majority of causal variants in the non-coding genomic regions.

Here, by leveraging large-scale genetic and observational data from public GWASs and UK Biobank (UKBB), we systematically investigated the genetic sharing patterns and epidemiological associations of three age-related ocular diseases including AMD, cataract, and glaucoma. We found extensive genetic pleiotropy and pairwise connections among these three diseases and identified seven pleiotropic loci across the genome. To better understand the mechanism underlying these pleiotropic loci, the integration of eye-specific gene eQTLs, epigenomic profiling, and 3D genome data were conducted to probe causal pleiotropic genes. Finally, drug repurposing was conducted by utilizing molecular networks for potential treatment or prevention of ocular comorbidities.

## Material and methods

### GWAS summary statistics collection

Full GWAS summary statistics of European ancestry on AMD, cataract, and glaucoma with the largest sample size were obtained after searching eight databases (GWAS Catalog ^24^, UKBB ^25^, GeneATLAS ^26^, GWAS Atlas ^27^, LD Hub ^28^, GRASP ^29^, PhenoScanner ^30^, and dbGaP ^31^). GWAS summary statistics with available signed statistics (beta or odds ratio), effect alleles, and non-effect alleles were retained during the search. We also collected GWAS summary data from FinnGen for replication analysis ^32^. More detailed information regarding the selected GWAS summary statistics for AMD, cataract, and glaucoma was shown in **Table S1**.

### Estimation of genetic correlation

Using GWAS summary statistics derived from the above datasets with a sample size > 50,000, we conducted LDSC ^33^ for these ocular diseases to estimate pairwise genetic correlations. SNPs were filtered to HapMap3 SNPs to avoid bias due to inconsistent imputation quality. SNPs with INFO ≤ 0.9, minor allele frequency ≤ 0.01, non-SNP variants, strand-ambiguous SNPs, and SNPs with duplicate IDs were removed. Pre-computed linkage disequilibrium (LD) scores and regression weights for European populations of the 1000 Genomes project were downloaded from the LDSC website (http://www.broadinstitute.org/~bulik/eur_ldscores/). We ensured that the LD score regression intercepts for AMD, cataract, and glaucoma were close to 1 during estimations. Moreover, genetic correlations between these diseases were also estimated using another tool SumHer from LDAK ^34^ under the recommended LDAK-Thin model. A tagging file computed using 2,000 white British individual genetic data files was downloaded from the LDAK website (https://dougspeed.com/pre-computed-tagging-files/), and the cutoff was set to 0.01 to preclude large-effect loci that explain at least 1% of phenotypic variance.

### Mendelian randomization

Mendelian randomization (MR) analysis between two traits was performed to determine whether genetic correlations were due to a causal link or confounding factors using the R package TwoSampleMR ^35^ which encompasses five MR methods (MR Egger, Weighted median, Inverse variance weighted, Simple mode, Weighted mode). The instruments for each exposure trait were significant SNPs (P-value < 5 × 10^−8^) after clumping, while the instruments for each outcome trait were all SNPs. Variants without SNP ID were annotated using the dbSNP ^36^ database; otherwise, SNPs were removed during subsequent analyses.

### Epidemiological analyses based on UK Biobank cohort

The UKBB is a large-scale cohort that recruited half a million UK participants (aged 37–73 years) between 2006 and 2010, and was followed up to 2021. Among the 502,412 participants, we excluded those who were pregnant (n = 379), lost to follow-up (n = 1,298), and had a mismatch between self-reported sex and genetic sex (n = 371), or missing information concerning covariates at baseline (n = 58,637). Overall, 441,727 participants were included in the study. AMD (ICD10: H35.3; ICD9: 362.5), cataract (ICD10: H25-H28; ICD9: 366), and glaucoma (ICD10: H40.1, H40.8, H40.9; ICD9: 365.1, 365.9) were ascertained based on information from self-reports, primary care records, hospital admissions, and death records. Multivariate Cox regression with separate models for incident AMD, cataract, and glaucoma was used to evaluate pairwise associations among them. In subsequent analyses, multi-state models were used to examine the role of one of the three ocular diseases in the transition between the start of follow-up and the other two disorders. In all models, we adjusted for sex, age, ethnicity, Townsend deprivation index, education level, smoking status, drinking status, physical activity, screen time-based sedentary behavior, diabetes, and other eye problems. This research was conducted using the UKBB resource under application number 83974.

### Determination of pleiotropic regions

We performed pleiotropic region analysis using gwas-pw ^37^, a tool for jointly analyzing pairs of GWAS traits. Overlapping SNPs of both traits were split into multiple blocks based on the bed files available at https://bitbucket.org/nygcresearch/ldetect-data. Separately, gwas-pw reckoned the posterior probability of the four models that the pleiotropic region 1) only associated with phenotype 1, 2) only associated with phenotype 2, 3) associated with both phenotypes, and 4) associated with both phenotypes independently.

### Identification of pleiotropic loci

To identify pleiotropic variants and characterize pleiotropy at single-variant level, cross-trait meta-analyses were carried out with GWAS summary statistics of three ocular diseases using three different approaches: 1) ASSET ^38^, 2) METAL ^39^, and 3) pleioFDR ^40^. ASSET explored all possible subsets of traits and evaluated a fixed-effect meta-analysis for each subset. The default scheme was selected when executing METAL with the correction of sample overlap. pleioFDR leveraged overlapping SNP associations of related phenotypes to identify genetic pleiotropic loci. All significant SNPs involving two or more traits were retained for the following analysis. Once classified, we conducted variant clumping through PLINK ^41^ and merged the pleiotropic loci if they physically overlapped (±250kb). The summary statistics of these diseases were standardized using a downloaded reference derived from genome files for 22 autosomes of filtered the 1000 Genomes project ^42^ European ancestry samples.

### Causal cell type estimation

We applied MAGMA ^43^ to evaluate whether specific ocular cell types showed significant heritability enrichment for traits based on ASSET one-sided summary statistics. MAGMA also requires gene average expression level across cell types derived from single-cell RNA sequencing and single-cell multi-omics sequencing data of anterior chamber angle or retina/choroid ^20, 21, 23, 44-46^ (**Table S2**). We additionally downloaded the raw data and determined the average expression values per cell type for each dataset using Cell Ranger ^47^ and the R package Seurat ^48^ unless the information was available. Cell types were assigned with the gene marker mentioned in the original publications, and the same cell types were manually renamed to maintain concordance with different single cell RNA-seq data. Gene names were transferred to Entrez gene IDs using the R package biomaRt ^49^. We appointed the direction of testing to “greater” and selected the condition-hide modifier for cell-type trait association analysis. We then leveraged the open chromatin peaks of retinal cell types from single-cell multiome to estimate LD scores for LDSC and conducted an additional cell-type trait association analysis.

### Causal SNP identification at pleiotropic loci

Colocalization analysis between two-eye eQTL data ^19, 50^ and ASSET one-sided GWAS data was conducted using the R package COLOC ^51^. We selected SNPs with an H4 posterior probability > 0.75, GWAS P-value < 1 × 10^−6^, and eQTL P-value < 0.05 as pleiotropic loci. By utilizing three sets of retina sample Assay of Transposase Accessible Chromatin sequencing (ATAC-seq) and H3K27ac Chromatin immunoprecipitation followed by sequencing (ChIP-seq) data ^52^, genomic enhancer regions of the retina were predicted using the ABC model ^53^ with default threshold settings. Next, we profiled the causal SNPs through fine-mapping analysis of pleiotropic loci identified by the above three meta-analysis methods. The one-sided ASSET meta-analysis results of pleiotropic SNPs located in the ± 1 Mb region of lead SNPs were imported into PolyFun ^54^ for fine-mapping with an embedded statistical method termed Susie ^55^, and 95% credible sets were revealed, assuming one causal SNP per locus. SNPs were included in the credible sets until the sum of the posterior probabilities was > 0.95.

### Pleiotropic causal gene prioritization

The causal genes in a pleiotropic locus were confined to the following rules: 1) Does the locus contain eye eQTL SNPs? If yes, eQTL harboring genes (eGenes) were selected. 2) Is the locus located in the open chromatin peaks from retina H3K27ac HiChIP data ^23^? If yes, causal genes were anchor-targeted genes. 3) Does the locus have a genomic region overlapping with the ABC model enhancer regions or a cell type-specific peak? If so, the causal genes were enhancer target genes or peak-inclusive genes. 4) If the pleiotropic locus contained none of the eQTL SNPs or was not mapped within any enhancer regions or peaks, the nearest gene with the highest fine-mapping posterior probability was identified as a causal gene. Moreover, gene ontology (GO) pathway enrichment analysis of causal genes with no more than 10 interactors was conducted in STRING ^56^ to shed light on pathogenic biological processes, then gene-phenotype association analysis and cell-type-specific enrichment analysis was conducted using Enrichr ^57^ and WebCSEA ^58^.

### Drug repurposing analysis

We curated approved drugs targeted at pleiotropic causal genes and target genes of such drugs for AMD, cataract, and glaucoma from Drugbank ^59^ and DGIdb ^60^ to verify whether the risk genes were identical to the drug target genes. Protein-protein interaction (PPI) network information was obtained from STRING and depicted using Cytoscape ^61^ to evaluate potential relationships between pleiotropic causal genes and drug target genes.

## Results

### Genetic sharing among AMD, cataract, and glaucoma

To delineate the shared genetic architecture among the three age-related ocular disorders, we first collected full GWAS summary statistics of AMD, cataract, and glaucoma from the latest public resources, incorporating three AMD GWASs, three cataract GWASs and two glaucoma GWASs, and most of them had sample size over 100,000 individuals (**Table S1**). Considering the signal overlaps of the three largest ocular GWAS summary statistics (**Figure S1A**), genetic correlations were measured using LD Score regression (LDSC) ^33^ and SumHer LDAK-Thin ^34^ models. Following careful quality control, we observed moderate and concordant genome-wide genetic correlations among these traits (**Figure 1A, 1B, Figure S1B**), in which glaucoma-cataract and AMD-cataract showed positive genetic correlations, while AMD-glaucoma exhibited negative genetic correlations. This is consistent with a previous study wherein the negative genetic correlation between AMD and glaucoma was reported ^16^. In addition, multiple bidirectional Mendelian randomization (MR) methods were applied to each pair of diseases to assess whether shared genetic components were coherent with true pleiotropy or mediated pleiotropy. Presuming that consistent results in three of the five MR methods indicate a strong causal effect between the two traits, there was no or weak evidence for potentially causal relationships among AMD, cataract, and glaucoma (**Figure S2**).

**Figure 1.**
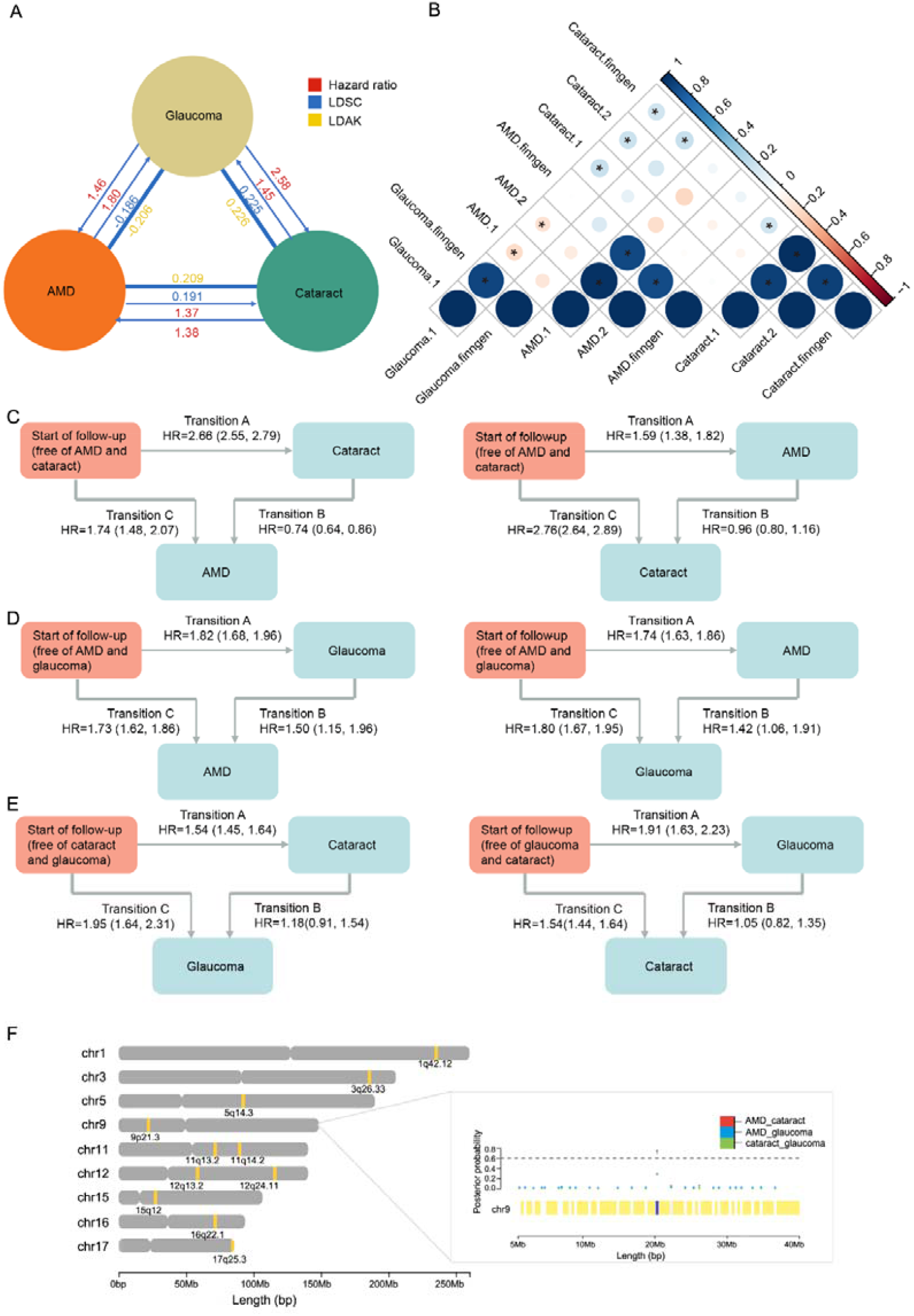
Genetic pleiotropy and epidemiological relationship of the three age-related ocular disorders. **A**. Epidemiological and genetic correlations between AMD, cataract, and glaucoma. Significant and strongest genetic correlations of disparate methods are shown. Red: hazard ratios estimated by Cox regression; blue: genetic correlations estimated by LDSC; yellow: genetic correlations estimated by LDAK. **B**. Genetic correlations of AMD, cataract, and glaucoma estimated by LDSC. The size and color of circles show the genetic correlations of disease pairs and the black stars indicate significant results. **C**. Multi-state models for the role of glaucoma in transitions to other two diseases. **D**. Multi-state models for the role of cataract in transitions to other two diseases. **E**. Multi-state models for the role of AMD in transitions to other two diseases. **F**. Chromosome graphics showing the pleiotropic regions assessed by gwas-pw. A partial enlarged view shows the pleiotropic region resided in chromosome 9 which overlapped with one locus identified by meta-analysis. Different color dots represent the posterior probability of trait pairs (red: AMD-cataract; blue: AMD-glaucoma; green: cataract-glaucoma). The dashed line represents the predictive threshold of posterior probability > 0.6.

### Epidemiological associations among AMD, cataract, and glaucoma

To test whether the pattern of genetic sharing for the three age-related ocular disorders is supported by epidemiological associations, we leveraged 441,727 selected participants from the UKBB with rich information regarding self-reports, primary care records, hospital admissions, and death records. In the UKBB cohort, at the end of follow-up, 36,5471 (82.74%) participants had no AMD, cataract, or glaucoma; 5,611 (1.27 %) had AMD alone; 48,639 (11.01%) had cataract alone; 7,431 (1.68%) had glaucoma alone; 8,251 (1.87%) had both AMD and cataract; 1,329 (0.30%) had both AMD and glaucoma; 7,015 (1.59%) had both cataract and glaucoma; and 1,010 (0.23%) had AMD, cataract, and glaucoma. Hazard ratios (HRs) estimated by Cox regression were significantly larger than 1 (P-value < 0.001), which indicated positive correlations among these traits (**Figure 1A, Figure S3**). These results showed significant pairwise associations between AMD, cataract, and glaucoma. Similar results were observed in further analyses using multi-state models which also uncovered positive correlations between cataract and other diseases as genetic correlation analysis showed. The correlation between AMD and glaucoma was inconsistent with our genetic correlation results which may due to environmental effects or inadequate power.

Additionally, multi-state models distinguished the roles of one of the three prevalent ocular diseases in the temporal trajectories of the other two. Prevalent glaucoma was inversely associated with AMD after cataract with HR of 0.74 (95% CI: 0.64-0.86), but not with cataract after AMD (**Figure 1C**). Multi-state analyses also showed that the associations of prevalent cataracts with the transition from the start of follow-up to subsequent AMD or subsequent glaucoma were stronger than those with the transition from AMD to glaucoma or from glaucoma to AMD (**Figure 1D**). Accounting for prevalent AMD in transitions of cataract and glaucoma, there was no evidence of stronger associations between prevalent AMD and transitions to cataract after having had glaucoma, or glaucoma after having had cataract (**Figure 1E**). Taken together, these new findings based on genetic evidence and epidemiological associations suggested a latent correlation among age-related ocular traits.

### Cross-disorder meta-analysis reveals seven pleiotropic loci

Considering the indication for genetic relationships between AMD-cataract, AMD-glaucoma, and cataract-glaucoma, 11 likely pleiotropic regions (posterior probability associated with two phenotypes > 0.6) were identified in the disease pairs using gwas-pw ^37^, one of which (chr9:20464018-2220526) turned out to be significant of two disease pairs (**Figure 1F, Table S3**). Next, we leveraged the largest collected GWAS summary statistics for AMD, cataract, and glaucoma containing 33,287 cases and 784,851 controls of European ancestry in total (AMD: 14,034 cases and 91,214 controls, cataract: 11,306 cases and 349,888 controls, glaucoma: 7,947 cases and 343,749 controls), and conducted cross-trait meta-analyses using three complementary methods including ASSET ^38^, METAL ^39^ and pleioFDR ^40^. Only those SNPs that surpassed genome-wide significance (P-value < 5 × 10^−8^) and had influences on at least two traits were reserved. For ASSET, two patterns were supported, and both were performed such that the one-sided ASSET meta-analysis maximized the standard fixed-effect meta-analysis test statistics over all possible subsets and guaranteed the identification of pleiotropic loci that have associations in the same direction, while the two-sided ASSET meta-analysis was performed in two directions automatically. Accordingly, we identified 9, 28, 6, and 7 pleiotropic loci using one-sided ASSET, two-sided ASSET, METAL, and pleioFDR, respectively, under the condition that the genomic inflation factor did not considerably deviate from one (**Figure 2A, Figure S4, Figure S5**).

**Figure 2.**
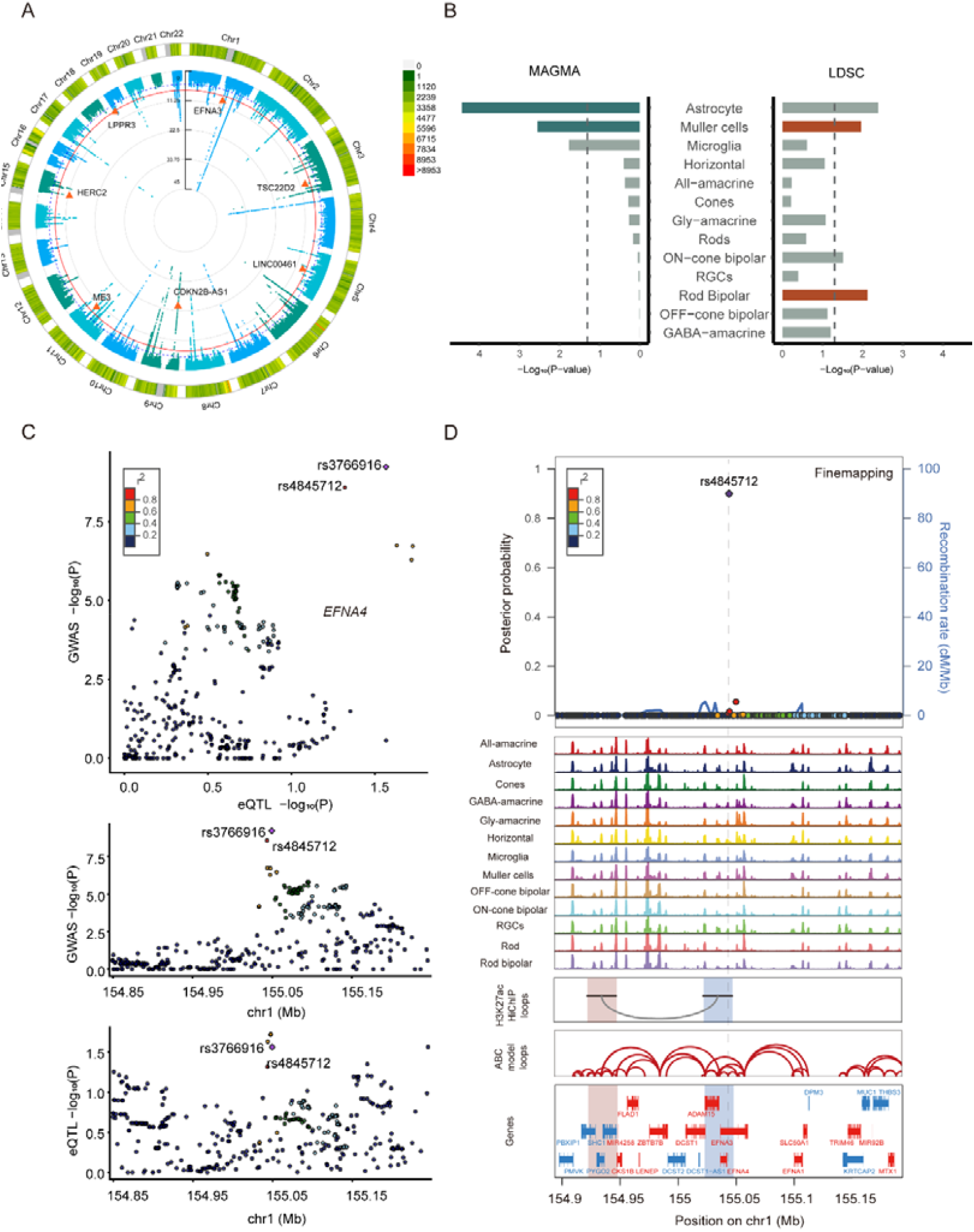
The pleiotropic loci of ocular phenotypes as well as trait-associated cell types. **A**. Circular Manhattan plot of the meta-analysis results using one-sided ASSET. The orange triangles denote the lead SNPs of pleiotropic loci identified by three meta-analysis methods. The texts beside the triangles show the nearest genes of these loci. The arcs in the outer layer represent chromosome positions and the number of SNPs in each position is depicted with different colors. Each dot in the inner layer represents a SNP. The red and blue circles indicate the genome-wide significant threshold (P-value□=□5 × 10^−8^) and nominal significant threshold (P-value□=□1 × 10^−5^). **B**. The significance of various cell types from MAGMA and LDSC in the cell-type trait association analysis. The significant cell types after Benjamini-Hochberg correction (FDR < 0.05) are highlighted. The dashed line indicates the threshold of P-value = 0.05. **C**. Locus-compare plots of lead SNP rs3766916 with macular eQTL gene *EFNA4*. Different colors of dots represent the linkage disequilibrium between the lead SNP (colored purple) and corresponding SNP. **D**. Locuszoom plots of the posterior probability of causality for variants around risk SNP rs4845712 which overlaps with *EFNA3* enhancer and retinal H3K27ac HiChIP data. The risk variant is uniquely highlighted with purple and all other SNPs are highlighted by their r^2^. The overlapped open chromatin peaks of 13 retinal cell types are shown in the middle. Anchor regions of the retina are highlighted with an orange or blue rectangle and red arcs show the enhance-gene pairs predicted by Activity-by-Contact model. Genes in red and blue represent the sense and antisense directions, respectively. The dashed line indicated the interested SNP position.

By guaranteeing that the lead SNPs should be marginally significant in all primary GWAS summary statistics and excluding the loci in the MHC region, seven loci were conservatively prioritized as candidate pleiotropic loci for which the one-sided ASSET results overlapped with the other three approaches (**Table 1**). Among these pleiotropic loci, most signals were significantly associated with glaucoma and were even stronger after meta-analysis although they were weaker in other diseases. Notably, three novel loci (lead SNPs: rs3766916, rs17421627, and rs146447071) became significant as the sample size increased. Besides, we found that three candidate pleiotropic loci intersected the aforementioned pleiotropic regions identified by gwas-pw (chr5:87390784-88891173, chr9:20464018-2220526, and chr15:110336875-113261782) (**Table 1, Figure 1F**), which rendered our results convincing. Furthermore, we collected additional GWAS datasets of AMD, cataract, and glaucoma from FinnGen project ^32^ and conducted a meta-analysis with ASSET, three loci (lead SNPs: rs4451405, rs10501626, and rs1129038) were replicated if the P-value of any SNPs in the same loci was smaller than 1 × 10^−5^ in one-sided ASSET or two-sided ASSET results.

**Table 1.** The seven pleiotropic loci identified by cross-disease meta-analysis approaches.

| Chr | SNP ID | Position | *P*-value (ASSET) | A1 | A2 | Diseases | Nearest gene(s) |
| --- | --- | --- | --- | --- | --- | --- | --- |
| 1 | rs3766916 | 155049402 | 5.78E-10 | T | C | AMD, cataract, glaucoma | *EFNA3* |
| 3 | rs35050931 | 150058352 | 5.22E-10 | T | A | cataract, glaucoma | *TSC22D2* |
| 5 | rs17421627 | 87847586 | 2.46E-09 | T | G | AMD, glaucoma | *LINC00461* |
| 9 | rs4451405 | 22071750 | 6.03E-25 | C | T | cataract, glaucoma | *CDKN2B-AS1* |
| 11 | rs10501626 | 86350791 | 2.04E-10 | G | T | AMD, cataract, glaucoma | *ME3* |
| 15 | rs1129038 | 28356859 | 1.84E-12 | C | T | AMD, cataract, glaucoma | *HERC2* |
| 19 | rs146447071 | 818751 | 2.52E-08 | T | C | cataract, glaucoma | *LPPR3* |

### Single-cell multi-omics data integration identifies likely causal cell types of ocular comorbidity

To understand the causal disease mechanisms underlying these pleiotropic loci, we investigated which cell types are essential during the development of ocular comorbidity. We obtained six single-cell RNA sequencing datasets and a single-cell multiome dataset derived from 33 human eyes **(Table S2)** and performed cell-type trait association analysis to explore the most likely causal cell types whose gene or epigenomic signatures correlate with genetic pleiotropy. In total, more than 40 cell types across the anterior chamber angle and retina/choroid were used in our study. Consequently, Müller cells and astrocytes were concordantly estimated to be two causal cell types of ocular disease susceptibility using both MAGMA and LDSC (**Figure 2B, Table S4, Table S5**), suggesting that the two cell types could play a critical role in modulating ocular comorbidity. Although astrocytes were not significant after Benjamini-Hochberg correction, it still showed trait-related using another macular scRNA dataset (**Figure S6**). In addition, vascular endothelium also displayed significant results in three MAGMA analyses (FDR < 0.05), which is consistent with previous study ^20^.

### Functional prioritization of causal pleiotropic variants and their target genes

To identify the causal genes acting on genetic pleiotropy across the three age-related ocular disorders, we first leveraged PolyFun ^54^ to fine-map the seven pleiotropic loci and identified causal variants in the 95% credible sets. We conducted colocalization analysis involving the seven pleiotropic loci based on two retinal *cis*-eQTL datasets ^19, 50^, yielding a credible variant rs3766916 colocalizing at several eGenes, including *EFNA4* and *TRIM46* (**Figure 2C, Figure S7, Table S6**). *TRIM46* encodes a protein of the tripartite motif family that controls neuronal polarity and high-glucose-induced ferroptosis in human retinal capillary endothelial cells ^62^, which suggests a potential causal role for retina-associated ocular traits.

Second, we collected retina H3K27ac HiChIP loops ^23^ to annotate all credible variants with spatially proximal genes. We found that two credible variants supported by HiChIP loops linked to different genes, and they were located in the eye enhancer regions or Müller cell/astrocyte open chromatin peaks according to single-cell multiome profiles. Notably, we found the SNP rs4845712 in high LD with rs3766916 (r^2^ = 0.95 in European) may be more likely to be the causal SNP at the corresponding pleiotropic locus. rs4845712 in the intron of gene *EFNA4* received the highest posterior probability of fine-mapping analysis and overlapped with a retina H3K27ac HiChIP loop (**Figure 2D**). rs4845712 host gene *EFNA4* has been reported to inhibit axonal regeneration and regulate axon guidance in the optic nerve ^63, 64^, and its HiChIP target gene *CKS1B* encodes a protein that binds to the catalytic subunit of cyclin-dependent kinases, and proliferation and migration of retinoblastoma cells can be inhibited by suppressing the expression of *CKS1B* ^65^. Additionally, we applied Activity-by-Contact (ABC) model ^53^ on ATAC-seq and H3K27ac ChIP-seq data from three retinal samples to predict enhancer-promoter interactions (**Table S7**), which indicated the SNP rs4845712 could interact with *EFNA3/EFNA4* gene enhancer.

Another top credible SNP rs1129038, close to *HERC2* promoter region, can interact with the genomic region containing *GABRA5* and *GABRB3* genes (**Figure 3A**). Among these genes, *HERC2* satisfied a genome-wide significance threshold with cataract in a multiethnic combined meta-analysis and was associated with uveal melanoma and macular inner retinal thickness ^66-68^. *GABRA5* and *GABRB3* coded GABA receptors that are important regulators of neuronal inhibition. Previous studies found SNPs within the *GABRB3* gene showed positive linkage with primary open-angle glaucoma and *GABRB3* disruption effectively reduced eye pigmentation resulting in visual defects in Angelman syndrome ^69, 70^. Based on the cell-type trait association analysis, target genes were achieved for an additional credible variant rs17421627. The SNP rs17421627 located at a pleiotropic locus was prioritized by three meta-analysis methods and obtained Müller cell/astrocyte-specific open chromatin signatures (**Figure 3B**). Target gene analysis based on cell-specific epigenomic profiling annotation suggested rs17421627 could interact with *LINC00461* gene. We found *LINC00461*, a gene that yields spliced long non-coding RNAs, was documented to be associated with several ocular traits, such as AMD, macular thickness, and macular telangiectasia type II ^71-73^. Finally, for other credible variants without evidence of active chromatin marks or chromosome interactions, we annotated the nearest gene as the target gene for the top credible variant at the pleiotropic locus. The annotated and prioritized causal variants and their genes were shown in **Table S8**.

**Figure 3.**
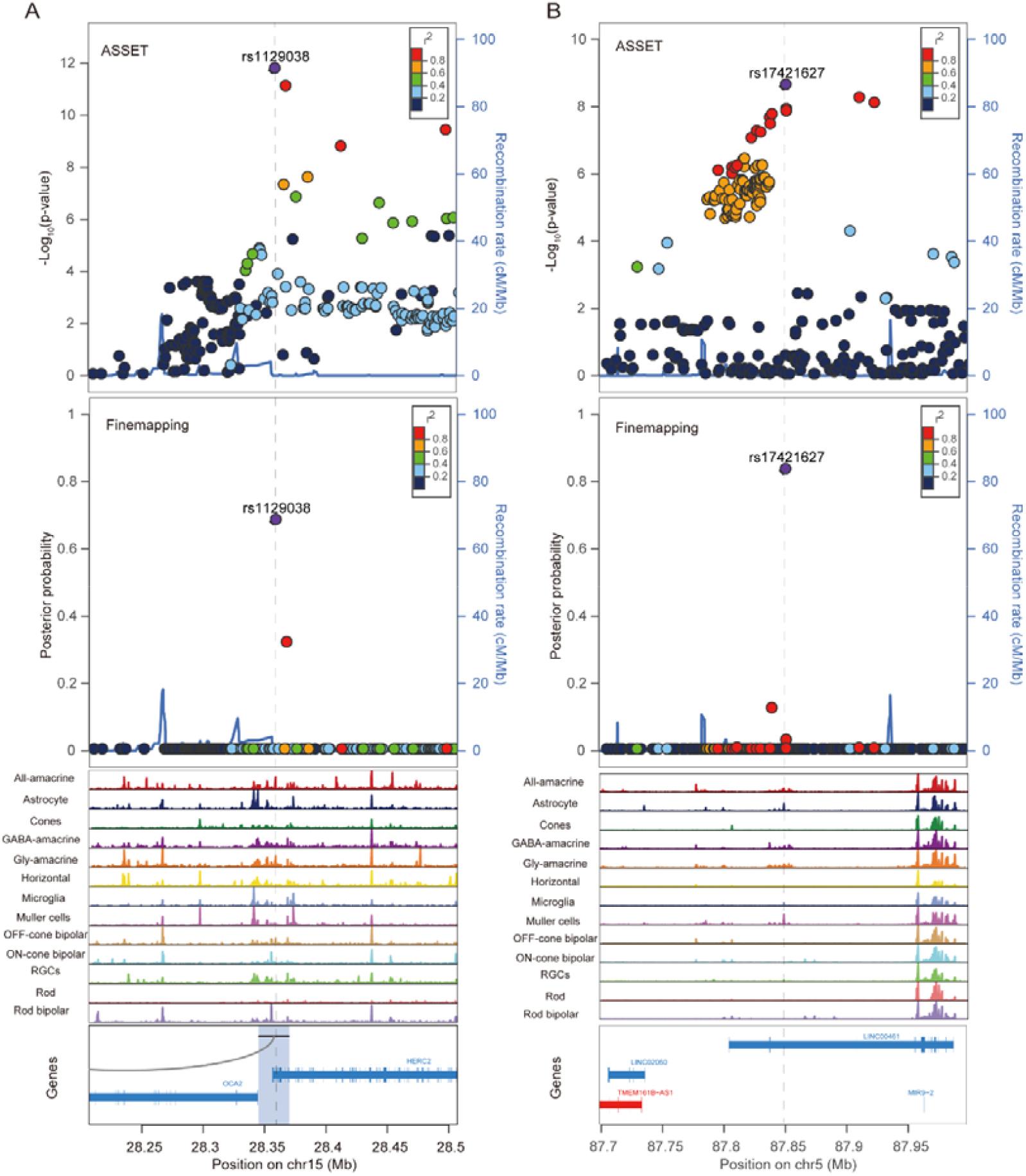
Integration with retinal eQTLs, epigenomic, and 3D genome data for causal genes prioritization. **A**. Locuszoom plots of one-sided ASSET associations and posterior probability of causality for variants around risk SNP rs1129038 which reside in one of the HiChIP anchors. **B**. Locuszoom plots of one-sided ASSET associations and posterior probability of causality for variants around risk SNP rs17421627 which overlaps with Müller cell/astrocyte-specific open chromatin peaks. The risk variant is uniquely highlighted with purple and all other SNPs are highlighted by their r^2^. The overlapped open chromatin peaks of 13 retinal cell types were shown in the middle. Anchor regions of the retina were highlighted with a blue rectangle. Genes in red and blue represent the sense and antisense directions, respectively. The dashed line indicated the interested SNP position.

### Shared biological pathways and drug repurposing opportunities

To assess the pleiotropic causal genes that affect biological pathways relevant to ocular diseases and explore the potential role of candidate genes in drug repurposing, we conducted several functional investigations. GO pathway analysis was first conducted in three orthogonal ontologies to explore the biological mechanisms of causal genes on ocular comorbidity. We detected 59 significant (FDR < 0.05) pathways, including the regulation of nerve development (GO:0048666, GO:0007409, GO:0021675, GO:0060080, GO:0007411, and GO:1904862) (**Figure 4A, Table S9**). Multiple glucose metabolic process pathways were also enriched, such as gluconeogenesis (GO:0006094), and pyruvate metabolic process (GO:0006090). We conducted gene-phenotype relationship analysis and revealed causal gene set was enriched for glaucoma as expected, macular thickness (q-value = 0.0293) and eye color (q-value = 0.0322). Cell-type-specific enrichment analysis showed causal genes were enriched in the visual cortex and inhibitory neurons (**Figure S8**). We then linked genes to licensed drugs and found aspirin and zopiclone targeted the causal genes *ME3* and *GABRA5* separately. By searching the approved drugs and drug targets for AMD, cataract, and glaucoma in Drugbank ^59^ and DGIdb ^60^, we identified 88 drug targets for the ocular diseases and found 6 (40%) of the 15 causal genes interacting with these drug targets. Noteworthily, *EFNA4*, a gene identified as being involved in neuronal development in previous pathway analysis, interacted with aflibercept-targeted gene *VEGFA* and pilocarpine-targeted gene *NTRK1* in the PPI network of AMD and glaucoma (**Figure 4B, Figure 4C**), indicating the drug repurposing opportunities for the treatment of ocular comorbidity.

**Figure 4.**
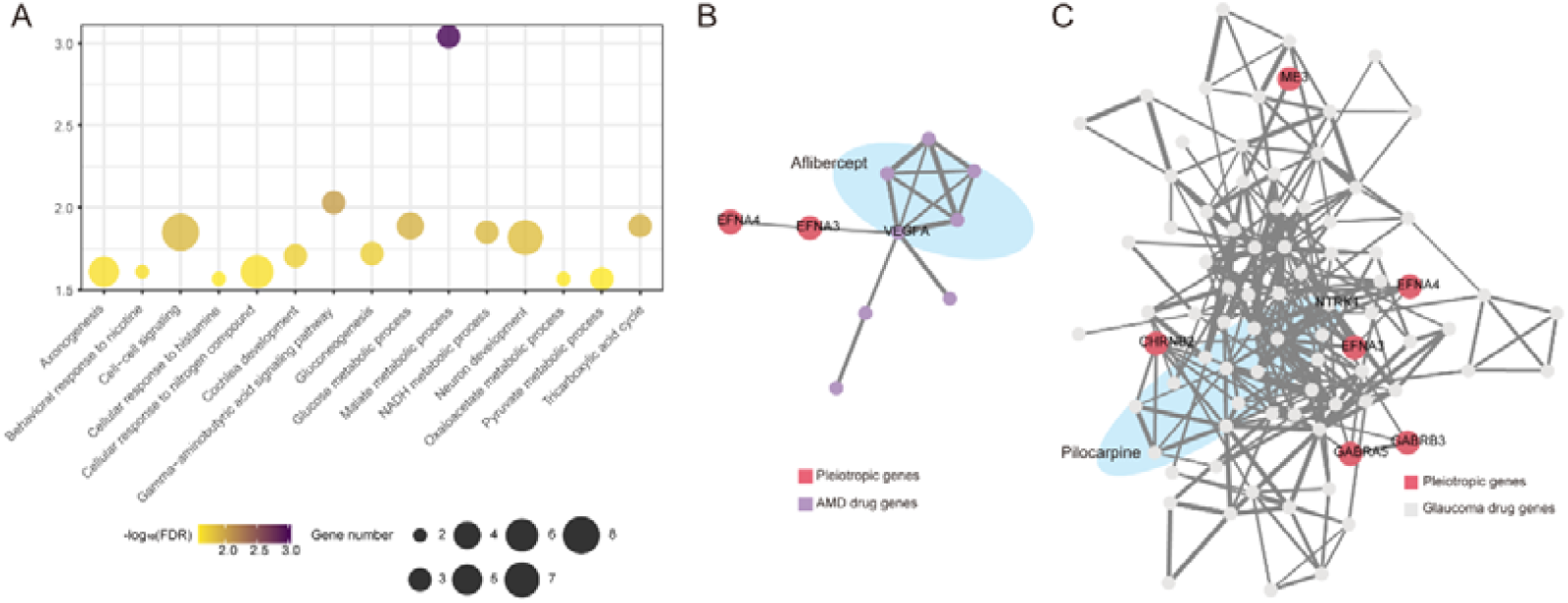
Functional enrichment of pleiotropic causal genes and drug repositioning for the treatment of ocular comorbidity. **A**. Gene ontology pathway enrichment analysis of pleiotropic causal genes. The size and color of the circles indicated the number of causal genes and the significance in the corresponding pathway. **B. & C**. Protein-protein interaction (PPI) networks using disease-risk genes (red) and drug-targeted genes (purple and light gray). The nodes in light blue ellipses represent the lateral drug target genes. The thickness of the edges represents the strength of the evidence.

## Discussion

In our study, we comprehensively investigated genetic sharing and epidemiological associations for three age-related ocular disorders. We confirmed the significant relationships among AMD, cataract, and glaucoma using a large-scale prospective observational study, genetic correlation analysis, and MR, followed by pleiotropic region analysis to explore regional pleiotropy. Seven significant pleiotropic loci of AMD, cataract, and glaucoma were identified using complementary meta-analyses, three of which achieved a marginally significant threshold in replication meta-analysis. In addition, we identified Müller cells and astrocytes as disease-associated cell types by single-cell multi-omics analysis. By incorporating statistical fine-mapping, retinal eQTL colocalization, and 3D genome loop data, we systematically prioritized causal target genes for pleiotropic variants. The interrogation of network biology was also implemented to identify disease genes-associated molecules and pathways for potential drug repositioning of ocular disorders.

To our knowledge, no previous study has investigated the genetic and non-genetic associations among AMD, cataract, and glaucoma. Several observational, clinical, and genetic evidence strongly suggest a shared biological basis or similar pathophysiological mechanisms among these age-related ocular disorders ^2-4, 16^. In this study, both epidemiological evidence based on large-scale UKBB data and genetic evidence based on several largest GWASs data supported pairwise tight associations between AMD, cataract, and glaucoma at different levels. The mechanisms behind our findings will shed light on the in-depth investigation of gene-environment interactions and comorbidity mechanisms across AMD, cataract, and glaucoma, which could also initiate further ocular disease comorbidity studies and emphasize the importance of comprehensive diagnosis and disease prevention in clinical practice.

By cross-disease meta-analysis at both variant and locus levels, we successfully identified seven pleiotropic loci. Nevertheless, understanding disease-causal cell types is of great value for the investigation of the precise mechanism between these loci-influenced genes and disease pathogenesis. We identified Müller cells and astrocytes as potential casual cell types for ocular comorbidity using two algorithms. A study claimed that Müller cells play a role in regulating serine biosynthesis, a critical part of the macular defense against oxidative stress, and their dysregulation may cause macular lesions ^74^. Furthermore, Müller cells and astrocytes have multiple functions symbiotically associated with RGCs (retinal ganglion cells), such as glutamate clearance, neurotrophic factor release, and neurotransmitter recycling ^75^, and the abnormality of RGCs will cause the impairment of visual acuity. Another study reported that the loss of astrocytes, together with retinal ischemia that occurs in the AMD macula, could induce the death of RGCs owing to oxidative damage ^76^. Despite insufficient evidence in our study, a connection between macular vascular endothelium and these ocular diseases has been widely discussed ^44, 77-80^.

Leveraging pleiotropic genes we found, we nominated several ocular disease-related pathways like the regulation of nerve development. The optic nerve is made up of RGC axons and transmits visual signals from photoreceptor cells to the brain. Diminution of vision will occur once the transitions were hindered. Glaucoma is a neurodegenerative disorder manifesting selective, progressive, and irreversible degeneration of the optic nerve, and optic nerve regeneration in glaucoma treatment was wildly studied ^81-83^. The photoreceptor synapses across the retinas of patients with AMD retract into the outer nuclear layer which evokes the subsequent outgrowth and reformation of synaptic contacts ^84^. Besides, optic nerve edema was found weeks to months after cataract extraction ^85^. By conducting gene-phenotype relationship analysis, pleiotropic genes were related to eye color, which suggested the relevance to these age-related ocular diseases. Darker iris color is pertinent with increased risks of cataract and intraocular pressure and mitigated risks of AMD, uveal melanoma, and pigmentary glaucoma ^14, 86-89^.

There are a few shortcomings in these analyses. First, the relationship between AMD and glaucoma was not consistent between genetic correlation analysis and epidemiological evidence which may be due to environmental impact factors. More studies should be conducted to clarify the relationship between AMD and glaucoma. Second, the number of glaucoma cases was much larger than those for the other two diseases, and some controls may inevitably overlap in glaucoma and cataract, which may affect the power of signals and cause systemic errors and wrongly preferential shared effects in meta-analysis. Third, in the present study, we found several cell types related to these traits, however, based on limited public annotation data, tissue-level annotation profiles were leveraged rather than cell type-specific data to identify truly causal genes. Thus, genetic information regarding the eye needs to be interpreted more precisely and accurately. Fourth, our fine-mapping only yielded credible sets of variants under the assumption of one causal variant; therefore, the prioritized causal genes may be underestimated. In addition, although we found several signs connecting certain drug targets and causal pleiotropic genes, whether these drugs are beneficial or harmful to the phenotype is still unknown. Further experiments concerning drug discovery and repurposing need to be conducted in the future.

## Data Availability

All data produced in the present work are contained in the manuscript

## Notes

**Funding information:** This study was supported by the National Natural Science Foundation of China [Grant Numbers 82020108007, 81830026, 32070675]; and the Beijing-Tianjin-Hebei Special Project [Grant Number 19JCZDJC64300(Z), 20JCZXJC00180].

### Competing Interest Statement

The authors have declared no competing interest.

### Funding Statement

National Natural Science Foundation of China (Grant Numbers 82020108007, 81830026, 32070675) and the Beijing-Tianjin-Hebei Special Project [Grant Number 19JCZDJC64300(Z), 20JCZXJC00180]

### Author Declarations

https://www.ebi.ac.uk/gwas/ http://www.nealelab.is/uk-biobank https://www.finngen.fi/en/access_results

### Summary of Updates

fix bugs because of sample size error

